# Group Testing with Homophily to Curb Epidemics with Asymptomatic Carriers

**DOI:** 10.1101/2020.10.09.20210260

**Authors:** Louis-Marie Harpedanne de Belleville

## Abstract

Contagion happens through heterogeneous interpersonal relations (homophily) which induce contamination clusters. Group testing is increasingly recognized as necessary to fight the asymptomatic transmission of the COVID-19. Still, it is plagued by false negatives. Homophily can be taken into account to design test pools that encompass potential contamination clusters. I show that this makes it possible to overcome the usual information-theoretic limits of group testing, which are based on an implicit homogeneity assumption. Even more interestingly, a multiple-step testing strategy combining this approach with advanced complementary exams for all individuals in pools identified as positive identifies asymptomatic carriers who would be missed even by costly exhaustive individual tests. Recent advances in group testing have brought large gains in efficiency, but within the bounds of the above cited information-theoretic limits, and without tackling the false negatives issue which is crucial for COVID-19. Homophily has been considered in the contagion literature already, but not in order to improve group testing.

## I. INTRODUCTION

Massive and timely identification of asymptomatic disease carriers is crucial if human-to-human asymptomatic transmission happens, which is documented for COVID19^1 2 3 4 5^. Li, Pei *et al*. (2020)^6^ find that although the transmission rate of undocumented carriers is only 55% that of documented carriers, the former are responsible for 80% of contaminations.

Massive and repeated RT-PCR testing is possible only through group testing (testing a pool of swabs of many individuals). Group testing is used to fight COVID19 in China, India, Germany, the United States^7^ and Rwanda^8^. For the literature on group testing and COVID19, see ^7 8 9 10 11 12 13 and 14^ With a .1 percent prevalence, the two-step adaptive design proposed by Dorfman (1943)^15^ decreases 17-fold the number of tests required to identify asymptomatic COVID-19 carriers (0.06 test per person) while the strategy suggested by Mutesa *et al*. (2020)^8^ decrease it 55 times (0.018 test per person). Still, these testing strategies are plagued by false negatives (see Section IV).

Contagion happens through heterogeneous interpersonal relations (homophily) which can be identified *ex ante* (Section II) to design test pools that encompass potential contamination clusters. Thus, it is possible to overcome information-theoretic limits, which rely on an implicit homogeneity assumption, and to make tests more efficient (Section III). Combining this approach with individual complementary exams for the positive groups identifies carriers who would be missed even by costly exhaustive individual tests on nasopharyngeal swabs (Section IV).

## II. HOMOPHILY IS PREVALENT AND CAN BE IDENTIFIED EX ANTE

The social sciences and epidemiology literatures show that heterogeneous interpersonal interactions, which induce small potential contamination clusters, are recurrent and can be identified *ex ante*. Thus, it is possible to design test pools encompassing potential clusters. Homophily, defined in 1954 by Lazarsfeld and Merton^16^, “refers to the fact that people are more prone to maintain relationships with people who are similar to themselves”^17^. Homophily is prevalent in many social networks^18^ and affects contagion^19^. Moulton (1986, 1990)^2021^, introduced a related econometric concept, clustering, which refers to the nondeterministic correlation of outcomes between individuals that are somewhat related: failing to take it into account induce significant errors when estimating standard errors. Clustering has been popularized by Bertrand *et al*. (2004)^22^ and extended to multiple non-nested dimensions^23^. Correcting for potential clustering is a condition *sine qua non* for publication in applied economics.

The epidemiologic literature confirms the importance of clusters. Han and Yang (2020)^24^ cite a Chinese-written article asserting that “In some cities, cases involving cluster transmission accounted for 50% to 80% of all confirmed cases of COVID-19.” According to a meta-analysis of 20 studies, households display high secondary attack rate (15.4% on average)^25^, and out of 36 children infected in a Chinese city, 32 (89%) had transmission by close contact with family members^26^. High SARs also occurs in a chalet (73.3%), at a choir (53.3%), at a religious event, or for travels and eating with an index case. Other cases of clusters with very high absolute attack rate include “a nursing home in Kings County, Washington (64%) […] a church in Arkansas (38%), a homeless shelter in Boston (36%), a fitness dance class (26.3%) and the Diamond Princess cruise ship in Japan (18.8%)”^25^. Park *et al*. (2020)^27^ analyze an outbreak in a building: 94 of 97 cases worked on the same floor, and 79 in the same open-space (attack rate of 52%). Many clusters have been observed in slaughterhouses.

*Ex ante* general contamination patterns in small clusters can be identified. Longer and more intense exposure increases the risk of infection^25 28 29 30^; so do indoor environments with sustained close contact and conversations^31^. In a call center, cases are concentrated in large open spaces but only one case in small offices^27^. This is consistent with Harpedanne (2020)^32^ who shows a convex theoretical contamination effect of the number of users. Using these general patterns and theoretical results, it is possible to identify *ex ante* potential clusters, and to design pools that encompass these clusters. Sections III and IV quantify the gains from this strategy.

## III. INFORMATION THEORETIC LIMITS CAN BE OVERCOME WHEN DESIGNING TEST POOLS

Here, I show that homophily contains information that can make group testing more efficient theoretically. For that purpose, I show that strong homophily used to design the testing pools makes it possible to overcome the most recent and tight information-theoretic lower bounds on the efficiency of group testing, identified by Chan *et al*. (2011)^33^ who claim to be the first in the literature to define limits in terms of actual numbers and not only rate or capacity, and Baldassini *et al*. (2013)^34^, who follow the same path and provide a new and tighter lower bound.

I focus here on noiseless tests: a negative test outcome is guaranteed when all items in the testing pool are nondefective, and a positive outcome when a least one item in the pool is defective^35^. Otherwise, the test is noisy.

Noisy tests are often examined under the assumptions of constant^33^ or worst-case^36^ noise. However, the literature points instead to a risk of false negatives increasing with dilution^14^ (analyzed in a previous draft), and to patient-specific idiosyncratic noise^37 38 39^. General form noise models (the symmetric error model^33^ or the additive model^40^) are irrelevant to analyze idiosyncratic noises. Also, focusing here on noiseless tests shows that the benefits of taking homophily into account in group testing are not limited to noise-related issues, and makes the comparison with the information-theoretic limit of Baldassini *et al*. (2013)^34^ easier. Furthermore, Baldassini *et al*. (2013, Section III)^34^ analyze a noiseless test in a population of size N, with K defectives (K is known for simplicity). They show that if the number of tests is limited to T, the probability of correct identification of the set of defectives is:

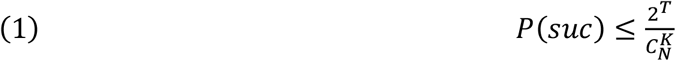

where 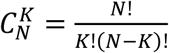.

Let build a simple counterexample. *N* is 64 and *K* is 8. Using 6 tests only (T=6), one can cut the population in 8 groups of 8 people each and determine which group contains carriers *if only one group contains carriers* (think of the 64 population as a 4×4×4 cube and cut it in half in each dimension, that is implement 6 tests over 32 people each). According to (1):

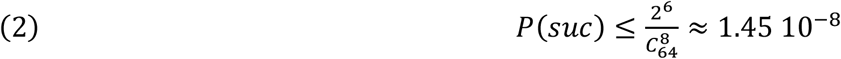

With homophily, there exists high potential for within-group contamination and low potential for between-groups contamination. With probability (1-ε_1_), only one individual has imported the disease in the 64 population (a decent assumption with a low general prevalence); ε_2_ is the probability that intergroup contamination has happened. Then with probability (1 - ε_1_)(1 - ε_2_), all 8 carriers are in the same group. Let fix ε_1_=0.2 and ε_2_=.5. Then:

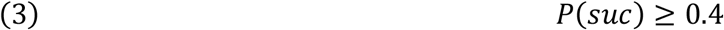

which contradicts (2): homophily provides information that makes it possible to overcome information-theoretic limits based on an implicit homogeneity assumption.

## IV. FALSE NEGATIVES CAN BE IDENTIFIED USING A TWO-STEP STRATEGY

The swabs of many disease carriers fail to contain viral loading, inducing patient-specific idiosyncratic false negatives^37 38 39^. A strategy based on group testing with homophily can solve this issue and identify more asymptomatic carriers than exhaustive individual testing.

Methods to identify SARS-CoV-2 carriers include clinical diagnosis (not for asymptomatic carriers), chest radiograph and CT-scan (not very available), fibrobronchoscope brush biopsy and RT-PCR on bronchoalveolar lavage fluid (requires specific equipment and skilled operators), sputum (produced in only 28 % of the COVID cases^41^), feces swabs, nasal and throat swabs… Only nasal and throat swabs can be used easily for large scale asymptomatic testing. The former induce a limited rate of false negatives if implemented less than one week after the onset of the disease for symptomatic patients (table 1). Thus, a strategy based on nasal swabs should be repeated weekly to minimize the risk of false negatives.

**Table 1 :**
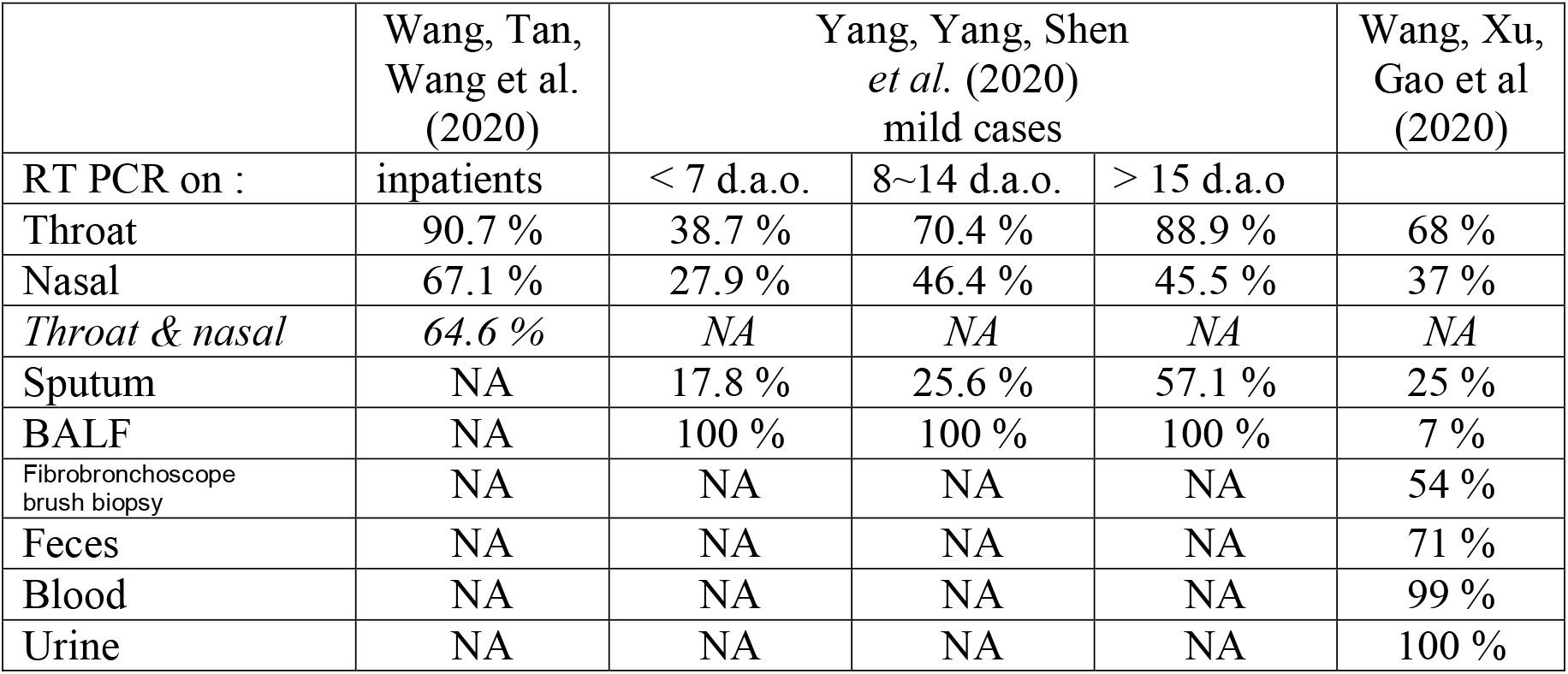
rate of false negatives on RT PCR tests

Still, even nasal swabs, widely used for identification of SARS-CoV-2, display a significant rate of false negatives. Group testing with homophily is very beneficial here. If a carrier is “false negative” with probability α, then a pool with two carriers will be negative with probability α^2^ only (α<1), and with probability *α*^*n*^ if there are *n* carriers: concentrating carriers in a pool decrease the risk of wrongly classifying this pool as negative.

In a second step, implementing individual RT-PCR on nasopharyngeal swab would reintroduce a false negative problem. Rather, advanced complementary exams are implemented on all individuals belonging to positive pools: chest radiograph or CT scan, fibrobronchoscope brush biopsy, and RT-PCR on additional types of swabs. With independence, the probability that all tests provide a false negative would be the product of the proportion of false negatives for each type of test. Empirical evidence on this issue is scarce: The correlation between nasal and throat swabs is low (Kappa=0.308) and Computed Tomography scan was always able to detect ground-glass opacities for cases without viral shedding in the swabs examined (3 cases)^39^. Similar results were obtained for PiO2/FiO2 and Murray score, implemented on two cases without viral shedding, always pointed to lung injury. Whether these results apply to asymptomatic carriers is an open question. More advanced work is needed on the correlation between these tests and exams for asymptomatic carriers, but a series of tests including nasal, throat and feces swabs, sputum swabs when available, lower respiratory swabs, PiO2/FiO2 tests and Murray score, may be able to detect most individual carriers if implemented frequently; CT scan may prove useful to identify more severe case.

Graph 1 analyze the quantitative gains from homophily in this two-step strategy, for 2 to 5 defectives. This covers a large range of situations: two defectives out of two thousand people correspond to a rate of 0.1%, while five out of 50 correspond to 10%. The size (and number) of pools do not affect the graphs, which therefore cover a large range of pool size.

From left to right, each graph displays increasing concentration (denoted by <C) of the defectives in testing pools. <_C_ is transitive but is not a total order, and using brackets ≈ for non-ordered configurations, we get, for 5 defectives:

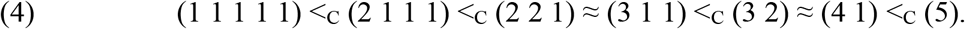

Bars describe the expected number of missed carriers due to false negatives. When carriers are concentrated in a few pools or a single pool, the expected number of missed carriers decreases. The gains from homophily can be summarized by comparing the expected number of missed carriers when all carriers are in different pools and when they are in the same pool. For *α*=1/3 (which is the central case^37 39^), the reduction reaches 67% for two carriers (0.67 expected missed carriers if the carriers are in two different groups against 0.222 if both are sin the same pool), 89% for three carriers, 96% for four carriers and 98.7% for five carriers. The absolute and relative gains are higher for *α*=0.5 (respectively 75%, 94%, 98.4% and 99.6% for two, three, four and five carriers) and lower for *α*=0.25 (respectively 50%, 75%, 87.5% and 93.75%). Even less successful concentration can still bring huge gains. For instance, with five carriers and *α*=1/3, getting three carriers in a pool and two in another brings a reduction of 80%.

**Graph 1:**
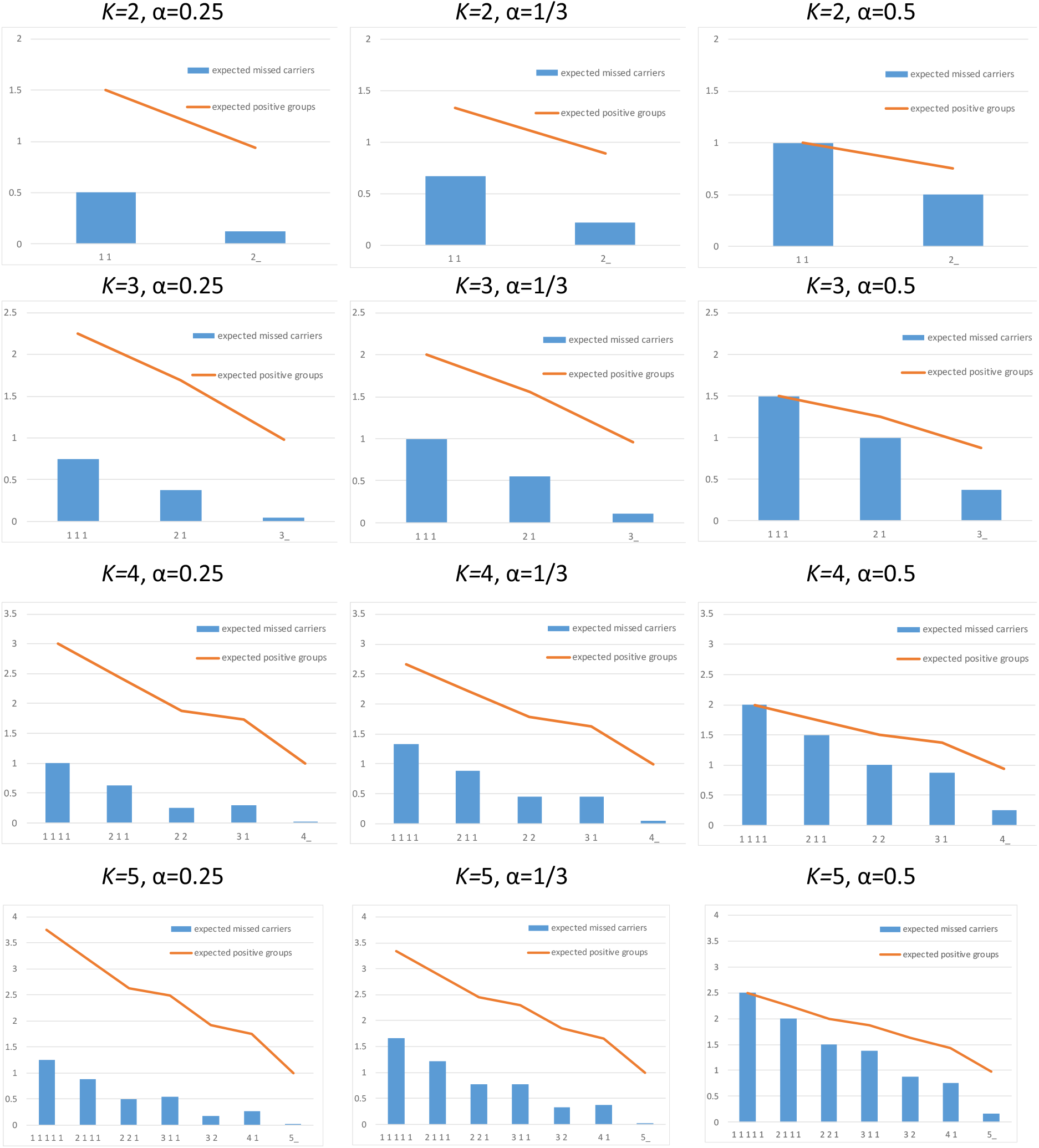
benefits of group testing and homophily to confront individual false negative

Note than even without homophily, that is if carriers are *i*.*i*.*d*. in the different pools, two carriers may be in the same pool by chance, which reduces the expected number of “missed” carriers. Group testing even without homophily can help identify and isolate carriers who would be missed by exhaustive individual testing. To the best of my knowledge, this simple and striking fact has been overlooked in the scientific and policy debates on group testing to fight epidemics. Advanced complementary exams are costly. Homophily is crucial here. When the concentration of carriers increases, the expected number of contaminated groups, and therefore the expected number of positive groups (groups that are *identified* as contaminated by the first step) are reduced stringently. The gains of homophily are lower than those observed for the expected number of missed carriers, because homophily has two opposite effects here: concentration decreases the number of potential positive groups, but improves the identification of these groups. Still, these gains are significant. For α=1/3, the reduction in the number of groups on which to implement complementary exams is 33.3% for two carriers, 51.8% for three carriers, 63% for four carriers and 70% for five carriers. The absolute gains are especially high for low values of α, since for low α, most contaminated groups are identified correctly and homophily reduces mainly the number of really contaminated groups.

## V. CONCLUSION

Homophily is prevalent in interpersonal interactions. Taking it into account makes group testing more efficient and help reduce false negatives issues. In case of a lockdown, testing together households, which are all potential clusters with high attack rate, is efficient; in more normal times, a multi-dimensional testing strategy with non-nested pools (households on the one hand, other potential cluster such as firms on the other hand) is likely to be beneficial. These results open a new avenue for research to fine tune the present analysis and combine it with other research in order to better fight the COVID19 epidemic.

The author declares no competing interest.

## Data Availability

No data: mathematical analysis

## Acknowledgements

I thank Marc Fleurbaey, Matthew Jackson, Xavier d’Haultfeuille and seminar participants at Paris School of Economics for useful comments. All remaining errors are mine.

## REFERENCES

1 Bai, Y., Yao, L., Wei, T., Tian, F., Jin, D.-Y., Chen, L., and Wang, M. (2020), “Presumed asymptomatic carrier transmission of COVID-19,” JAMA. (Epub ahead of print)

2 Rothe, C., Schunk, M., Sothmann, P., Bretzel, G., Froeschl, G., Wallrauch, C.,. Zimmer, T., Thiel, V., Janke, C., Guggemos, W., Seilmaier, M., Drosten, C., Vollmar, P., Zwirglmaier, K., Zange, S., Wölfel, R., Hoelscher, M., (2020, March 5), “Transmission of 2019-nCoV infection from an asymptomatic contact in Germany,” New England Journal of Medicine (Epub ahead of print)

3 Zou L., Ruan, F., Huang, M., Liang, L., Huang, H., Hong, Z., Yu, J., Kang, M., Song, Y., Xia, J., Guo, Q., Song, T., He, J., Yen, H. L., Peiris, M., and Wu, J. (2020, March 19), “SARS-CoV-2 viral load in upper respiratory specimens of infected patients,” New England Journal of Medicine.

4 Santarpia, J. L., Rivera, D. N., Herrera, V., Morwitzer, M. J., Creager, H., Santarpia, G. W., Crown, K. K., Brett-Major, D. M., Schnaubelt, E., Broadhurst, M. J., Lawler, J. V., St. Reid, P., and Lowe, J. J. (2020, March 26), “Transmission potential of SARS-CoV-2 in viral shedding observed at the University of Nebraska Medical Center,” mimeo.

5 Wong, Justin, Anita B Z Abdul Aziz, Liling Chaw, Abdirahman Mahamud, Matthew M. Griffith, Ying-Ru Lo, Lin Naing (2020) “High proportion of asymptomatic and presymptomatic COVID-19 infections in travelers and returning residents to Brunei”, Oxford University Press – Public Health Emergency Collection.

6 Li, Ruiyun, Sen Pei, Bin Chen, Yimeng Song, Toa zang, Wan Yang and Jeffrey Shaman (2020), “Substantial undocumented infection facilitates the rapid dissemination of novel coronavirus (SARS-CoV-2)”, Science, 368:489–493. doi:10.1126/science.abb3221

7 Mallapaty, Smriti (2020), “The mathematical strategy that could transform coronavirus testing”, Nature (10 July 2020). https://www.nature.com/articles/d41586-020-02053-6

8 Mutesa, Leon, Pacifique Ndishimye, Yvan Butera, Jacob Souopgui, Annette Uwineza, Robert Rutayisire, Ella Larissa Ndoricimpaye, Emile Musoni, Nadine Rujeni, Thierry Nyatanyi, Edouard Ntagwabira, Muhammed Semakula, Clarisse Musanabaganwa, Daniel Nyamwasa, Maurice Ndashimye, Eva Ujeneza, Ivan Emile Mwikarago, Claude Mambo Muvunyi, Jean Baptiste Mazarati, Sabin Nsanzimana, Neil Turok & Wilfred Ndifon (2020) “A pooled testing strategy for identifying SARS-CoV-2 at low prevalence”, Nature, https://www.nature.com/articles/s41586-020-2885-5

9 Gollier, Christian and Olivier Gossner (2020), “Group Testing against COVID-19”, CREST DT 2020-04.

10 Conger, K. (2020, April). Testing pooled samples for COVID-19 helps Stanford researchers track early viral spread in Bay Area. Stanford Medicine News Center.

11 Lakdawalla, Darius, Emmett Keeler, Dana Goldman, Erin Trish (2020), “Getting Americans Back to Work (and School) with Pooled Testing”, USC Schaeffer Center White Paper.

12 Eberhardt, Jens Niklas, Nikolas Peter Breuckmann and Christiane Sigrid Eberhardt (2020), “Multi-Stage Group Testing Improves Efficiency of Large-Scale COVID-19 Screening”, Journal of Clinical Virology doi: 10.1016/j.jcv.2020.104382

13 Lohse, Stefan,Thorsten Pfuhl, Barbara Berkó-Göttel, Jürgen Rissland, Tobias Geißler, Barbara Gärtner,., Sören L Becker, Sophie Schneitler, Sigrun Smola (2020, April 28), “Pooling of Samples for testing for SARS-CoV-2 in asymptomatic people,” Lancet Infectious Diseases. https://doi.org/10.1016/S1473-3099(20)30362-5.

14 Yelin, Idan, Noga Aharony, Einat Shaer Tamar, Amir Argoetti, Esther Messer, Dina Berenbaum, Einat Shafran, Areen Kuzli, Nagam Gandali, Tamar Hashimshony, Yael Mandel-Gutfreund, Michael Halberthal, Yuval Geffen, Moran Szwarcwort-Cohen and Roy Kishony (2020), “Evaluation of COVID-19 RT-qPCR test in multi-sample pools”, Clinical Infectious Diseases. doi: 10.1093/cid/ciaa531.

15 Dorfman, R. (1943), “The detection of defective members of large population”, The Annals of Mathematical Statistics, 436–440.

16 Lazarsfeld, Paul and Robert K. Merton (1954), “Friendship as a Social Process: A Substantive and Methodological Analysis”, pp. 18–66 in Freedom and Control in Modern Society, edited by M. Berger, T. Abel, and C. H. Page. New York: Van Nostrand.

17 Jackson, Matthew O. (2008), “Social and Economic Networks”, Princeton University Press, Princeton, New Jersey, 504 pp.

18 McPherson, Miller; Lynn Smith-Lovin and James M. Cook (2003). “Birds of a Feather: Homophily in Social Networks”, Annual Review of Sociology, Vol. 27 (1), pp. 415–444.

19 Jackson, M. O., and Lopez-Pintado, D. (2013), “Diffusion and contagion in networks with heterogeneous agents and homophily,” Network Science 1(1):49–67.

20 Moulton, Brent R. (1986), “Random Group Effects and the Precision of Regression Estimates”, Journal of Econometrics, 32: 385–397.

21 Moulton, Brent R. (1990), “An Illustration of a Pitfall in Estimating the Effects of Aggregate Variables on Micro Units”, Review of Economics and Statistics 72(3): 334–338.

22 Bertrand, Marianne, Esther Duflo and Sendhil Mullainathan (2004), “How Much Should We Trust Difference-in-Difference Estimates?”, The Quarterly Journal of Economics, Vol. 119 (1), pp. 249–275

23 Cameron, A.Colin., J. G. Gelbach and Douglas L. Miller (2011), “Robust Inference with Multi-Way Clustering”, Journal of Business and Economic Statistics 29(2): 238–249.

24 Han, Yu and Hailan Yang (2020), “The transmission and diagnosis of 2019 novel coronavirus infection disease (COVID-19): A Chinese perspective”, Journal of Medical Virology 92, 639–644.

25 Koh, Wee Chian L, in Naing, Muhammad Ali Rosledzana, Mohammad Fathi Alikhan, Liling Chaw, Matthew Griffith, Roberta Pastore and Justin Wong (2020), “What do we know about SARS-CoV-2 transmission? A systematic review and meta-analysis of the secondary attack rate, serial interval, and asymptomatic infection”, medRxiv.

26 Qiu H, J. Wu, L. Hong, Y. Luo, Q. Song and D. Chen (2020) « Clinical and epidemiological features of 36 children with coronavirus disease 2019 (COVID-19) in Zhejiang, China: an observational cohort study. Lancet Infectious Disease doi:10.1016/S1473-3099(20)30198-5

27 Park, Shin Young, Young-Man Kim, Seonju Yi, Sangeun Lee, Baeg-Ju Na, Chang Bo Kim, Jungil Kim, Hea Sook Kim, Young Bok Kim, Yoojin Park, In Sil Huh, Hye Kyung Kim, Hyung Jun Yoon, Hanaram Jang, Kyungnam Kim, Yeonhwa Chang, Inhye Kim, Hyeyoung Lee, Jin Gwack, Seong Sun Kim, Miyoung Kim, Sanghui Kweon, Young June Choe, Ok Park, Young Joon Park, Eun Kyeong Jeong (2020), “Coronavirus Disease Outbreak in Call Center, South Korea”, Emerging Infectious Diseases 26 (8).

28 Li W, Zhang B, Lu J, Liu S, Chang Z, Cao P, et al. (2020) “The characteristics of household transmission of COVID-19”. Clinical Infectious Disease; ciaa450.

29 Madewell, Zachary J., Yang Yang, Ira M. Longini Jr, M. Elizabeth Halloran, Natalie E. Dean “Household transmission of SARS-CoV-2: a systematic review and meta-analysis of secondary attack rate”, medRxiv.

30 Liu T, Liang W, Zhong H, et al. (2020) “Risk factors associated with COVID-19 infection: a retrospective cohort study based on contacts tracing”. Emerging Microbes & Infection: 1–31.

31 Nishiura H, Oshitani H, Kobayashi T, et al. (2020) “Closed environments facilitate secondary transmission of coronavirus disease 2019 (COVID-19)”. medRxiv.

32 Harpedanne de Belleville, Louis-Marie (2020), Act Now or Forever Hold Your Peace: Slowing contagion with Unknown Spreaders, Limited Cleaning Capacities, and Costless Measures, mimeo. https://papers.ssrn.com/sol3/papers.cfm?abstract_id=3562739 https://mpra.ub.uni-muenchen.de/99728/

33 Chan, Chun Lam, Pak Hou Che, Sidhart Jaggi and Venkatesh Saligrama (2011), “Non-adaptative probabilistic group testing with noisy measurments: Near-optimal bounds with efficient algorithms”, in 49th Allerton Conference, pp. 1832–1839.

34 Baldassini, Leonardo, Oliver Johnson and Matthew Aldridge (2013), “The Capacity of Adaptative Group Testing”, IEEE International Symposium on Information Theory, 2676-2680.

35 Aldridge, M, Johnson, O, and Scarlett, J. (2019, October 2), “Group Testing: An Information Theory Perspective,” Foundations and Trends in Communications and Information Theory 15(3-4): 196–392.

36 Macula, Anthony J. (1997) “Error-correcting nonadaptive group testing with dedisjunct matrices,” Discrete Applied Mathematics, vol. 80, no. 2-3, pp. 217–222, 1997.

37 Yang, Yang, Minghui Yang, Chenguang Shen, Fuxiang Wang, Jing Yuan, Jinxiu Li, Mingxia Zhang, Zhaoqin Wang, Li Xing, Jinli Wei, Ling Peng, Gary Wong, Haixia Zheng, Weibo Wu, Mingfeng Liao, Kai Feng, Jianming Li, Qianting Yang, Juanjuan Zhao, Zheng Zang Lei Liu and Yingxia Liu (2020, February). “Evaluating the accuracy of different respiratory specimens in the laboratory diagnosis and monitoring the viral shedding of 2019-nCoV infections”.

38 Wang, Wenling, Yanli Xu, Ruqin Gao, Roujian Lu, Kai Han, Ghuizen Wu and Wenjie Tan (2020, March). “Detection of SARS-CoV-2 in different types of clinical specimens” JAMA. doi:10.1001/jama.2020.3786.

39 Wang, Xiong, Li Tan, Xu Wang, Weiyong Liu, Yanjun Lu, Liming Cheng, Ziyong Sun (2020), “Comparison of nasopharyngeal and oropharyngeal swabs for SARS-CoV-2 detection in 353 patients received tests with both specimens simultaneously”, International Journal of Infectious Diseases 94, 107–109

40 Atia, G. and V. Saligrama, “Boolean compressed sensing and noisy group testing” IEEE Trans. Inform. Theory, vol. 58, no. 3, pp. 1880–1901, March 2012.

41 Huang C, Wang Y, Li X, et al. (2020), “Clinical features of patients infected with 2019 novel coronavirus in Wuhan, China”, Lancet.

